# Navigating primary care in Ontario: a qualitative study investigating the perceptions of Chinese newcomers to Canada

**DOI:** 10.64898/2025.12.01.25341083

**Authors:** Stephen Marisette, Joanne A. Permaul, Zhiheng Zeng, Junyi Mei, Donatus Mutasingwa, Melanie Henry, Andrea Groff, Kathleen Homiak

## Abstract

Newcomers to Canada often encounter challenges navigating a novel healthcare system. Some of these challenges may be related to differences between the healthcare system in their home country and the Canadian system. Few studies have addressed how Chinese newcomers to Canada understand the role of primary care; this study addresses this gap. The primary objective was to explore how Chinese newcomers perceive the role of primary care in Ontario. Secondary objectives explored how they learned about primary care, their understanding of continuity of care, and their perceptions of preventive healthcare. This qualitative study used individual interviews conducted with residents of Ontario who immigrated from mainland China in the last 5 years. Transcripts from 10 interviews were analyzed using thematic analysis and demographic data were analyzed descriptively. Main themes included: barriers to accessing care, differences between healthcare systems, the importance of continuity of care, understanding the family doctor’s role, the significance of preventive healthcare, and healthcare system information needs. Given the substantial differences in how healthcare is provided in China compared to Canada, there is a need to improve newcomer education and orientation regarding the role of primary care in Ontario. Improved understanding of the Canadian healthcare system would help newcomers address barriers and assist with system navigation. Study findings also have broader implications for understanding the experiences and perceptions of newcomers from other countries as they navigate primary care in a new setting.

## Background

Newcomers to Canada face multiple barriers to accessing healthcare including language barriers, cultural differences, socioeconomic challenges, and healthcare system factors such as the referral process and long wait times [1-10]. Analysis of the 2009/2010 Canadian Community Health Survey revealed that, compared to native-born Canadians and “established” immigrants, recent immigrants to Canada were more likely to use walk-in clinics and emergency rooms, and less likely to visit a family physician [9]. Canada’s primary care-based system has been identified as a key barrier to access for newcomers to Canada [5,6,9]. A qualitative study of Bangladeshi immigrants to Canada found that some participants were unaware that the Canadian healthcare system is structured around a model in which patients are intended to have a primary care provider [6].

The 2016 census data showed that in Markham, Ontario (the setting for this study), 58% of the population were immigrants to Canada; 11% of these newcomers had immigrated within the previous five years [11]. This study focused on newcomers from mainland China because, according to this census data, China was the country of birth for 31% of immigrants who reside in Markham [11]. Newcomers from China may face challenges navigating the Canadian healthcare system due to differences compared to the Chinese healthcare system. In Canada, publicly funded universal healthcare covers most medical services; however, non-emergency services can have long wait times due to a shortage of healthcare professionals, underfunding of primary care, and a high demand for services [12]. China has a combination of government-subsidized and private healthcare options which can lead to disparities in access to quality healthcare based on geographic location and the ability to pay for private services [13]. Although China has enacted reforms to strengthen primary care, results are limited in part by a payment system which incentivizes testing and treatment over prevention [14].

While the literature has identified limited familiarity with the Canadian healthcare system as a significant challenge for some immigrants, there remains an inadequate appreciation of how newcomers understand primary care in Canada. This study is the first to examine perceptions of primary care among newcomers from mainland China living in Ontario, Canada. Its primary objective was to explore how newcomers perceived the role of primary care in Ontario, with secondary objectives examining how they learned about the system, their understanding of continuity of care, and their perceptions of preventive healthcare.

### Theoretical Framework

This qualitative study operates within a constructivist paradigm. In contrast to positivism, constructivism asserts that there is no one objective reality that we all experience; rather, an individual’s background, interactions, and experiences shape their perspective of reality [15]. Thematic analysis, which aligns well with the constructivist paradigm [16], was used to analyze qualitative data in this study.

## Methods

### Participants

The Oak Valley Health Research Ethics Board approved this study. Participants were recruited by the Centre for Immigrant and Community Services (CICS) in Markham – a local community agency that helps newcomers to settle in Canada. Markham area residents, who were 25 years of age or older, had immigrated from mainland China in the last 5 years, and could communicate in English or Mandarin, were eligible to participate. Given the unique barriers to care faced by temporary foreign workers, refugees, and undocumented individuals, these groups were excluded from participating in the study.

### Data Collection

CICS staff invited eligible clients to participate by distributing the study flyer, and through CICS chat groups and social media accounts. Interested clients were given the consent form to review at home and invited to contact the study investigators for further information. The flyer and consent form were available in English and Chinese.

One-on-one, semi-structured, individual interviews were conducted in English or Mandarin, by one of two study investigators, who were fluent in both languages, between January and August 2023. Interviews were approximately one hour in length and conducted virtually, either by Zoom or telephone. Verbal consent was obtained by the interviewer prior to the interview, and a demographic questionnaire was administered. An interview guide was developed collaboratively by the study team to facilitate the interviews and to provide a framework for discussion (Table 1). Interviews were audio-recorded, transcribed, and translated, if required. Participants who completed the interview received a $40 grocery gift card to compensate them for their time.

**Table 1.**
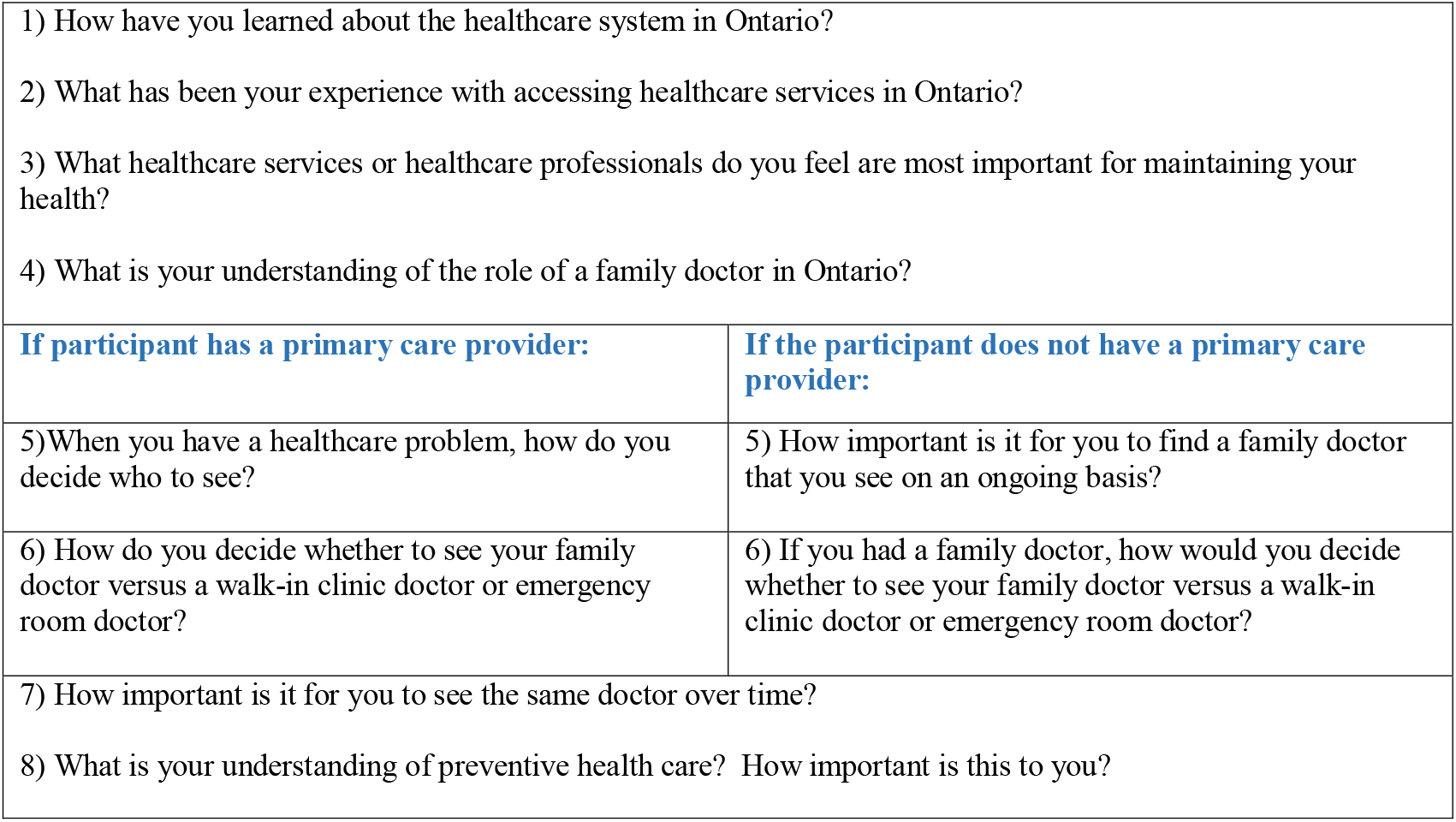
Questions from the Semi-Structured Interview Guide.

### Data Analysis

Data from the demographic questionnaires were analyzed descriptively using SPSS^©^ Version 28 to conduct frequency distributions and means analysis, and NVivo^©^ software was used to code the data from the transcripts. Analysis of data from the interviews was concurrent with ongoing data collection. Three researchers analyzed the transcripts using thematic analysis [17]. There were six steps associated with this process: familiarization with the data, generating initial codes, searching for themes, reviewing themes, defining and naming themes, and producing the final report [16]. Three members of the research team independently coded the interview transcripts and then convened to reach consensus on coding and to discuss emerging themes.

We approached data analysis from a constructivist standpoint, believing that the concept of data saturation, as traditionally defined, does not align with this paradigm [17]. Instead of striving for saturation, our goal was to achieve a deep and nuanced understanding of participants’ experiences, a process in which the researchers are active participants [18]. Generally, a study with a narrow aim and specific population requires fewer interviews to obtain this deeper understanding compared to a study with a broader aim and few inclusion/exclusion criteria [17]. It was anticipated that a sample size of 10 to 15 would provide an adequate understanding of how Chinese newcomers to the Markham area perceive primary care in Ontario.

Guidelines for qualitative research were followed to ensure rigour and trustworthiness of the data [19]: the interviewers were familiar with the culture of the participants; purposive sampling was used to select participants who met specific inclusion criteria; in addition to the interviews, a survey was used to collect demographic data; iterative questioning was used, including probes to elicit detailed responses; interviewers checked the accuracy of the transcription and translation of interviews; and an audit trail was maintained throughout the analysis.

## Results

Ten individuals participated in one-on-one interviews for this study, which were conducted by telephone or Zoom video conferencing. Participant demographics are described in Table 2. Although all participants spoke English, eight participants requested to have the interview conducted in Mandarin, and two in English. Six themes were derived from the coded data which are interrelated, as described in the thematic map (Figure 1).

**Table 2.** Participant Demographics (N=10)

|  |  |
| --- | --- |
| Age (mean, range): | 44 years, 30 – 68 years |
| Female (N, %): | 8, 80% |
| Married (N, %): | 9, 90% |
| Number of people in household (mean, range): | 4 people, 2 – 5 people |
| Time since arriving in Canada (mean, range): | 25 months, 1 – 58 months |
| Speak English (N, %): | 10, 100% |
| Completed college/university (N, %): | 9, 90% |
| Employed (N, %): | 7, 70% |
| Income (N, %):<br>0 – \$29,999<br>\$30,000 - \$59,999<br>\$60,000 - \$89,999<br>\$90,000 - \$119,999<br>\$120,000 - \$149,999<br>\$150,000 or more<br>Don't know<br>Prefer not to answer | 1, 10%<br>2, 20%<br>1, 10%<br>1, 10%<br>1, 10%<br>0<br>2, 20%<br>2, 20% |
| Income (N, %):<br>< \$60,000<br>≥ \$60,000<br>Don't know<br>Prefer not to answer | 3, 30%<br>3, 30%<br>2, 20%<br>2, 20% |
| Have someone to assist with navigating healthcare system (N, %) | 6, 60% |
| Have a primary care provider in Canada (N, %) | 8, 80% |
| Had a primary care provider before coming to Canada (N, %) | 1, 10% |
| Have a chronic illness (N, %)<br>(responses: hypertension, cardiovascular disease, migraine headaches, chronic gastritis) | 4, 40% |

**Figure 1.**
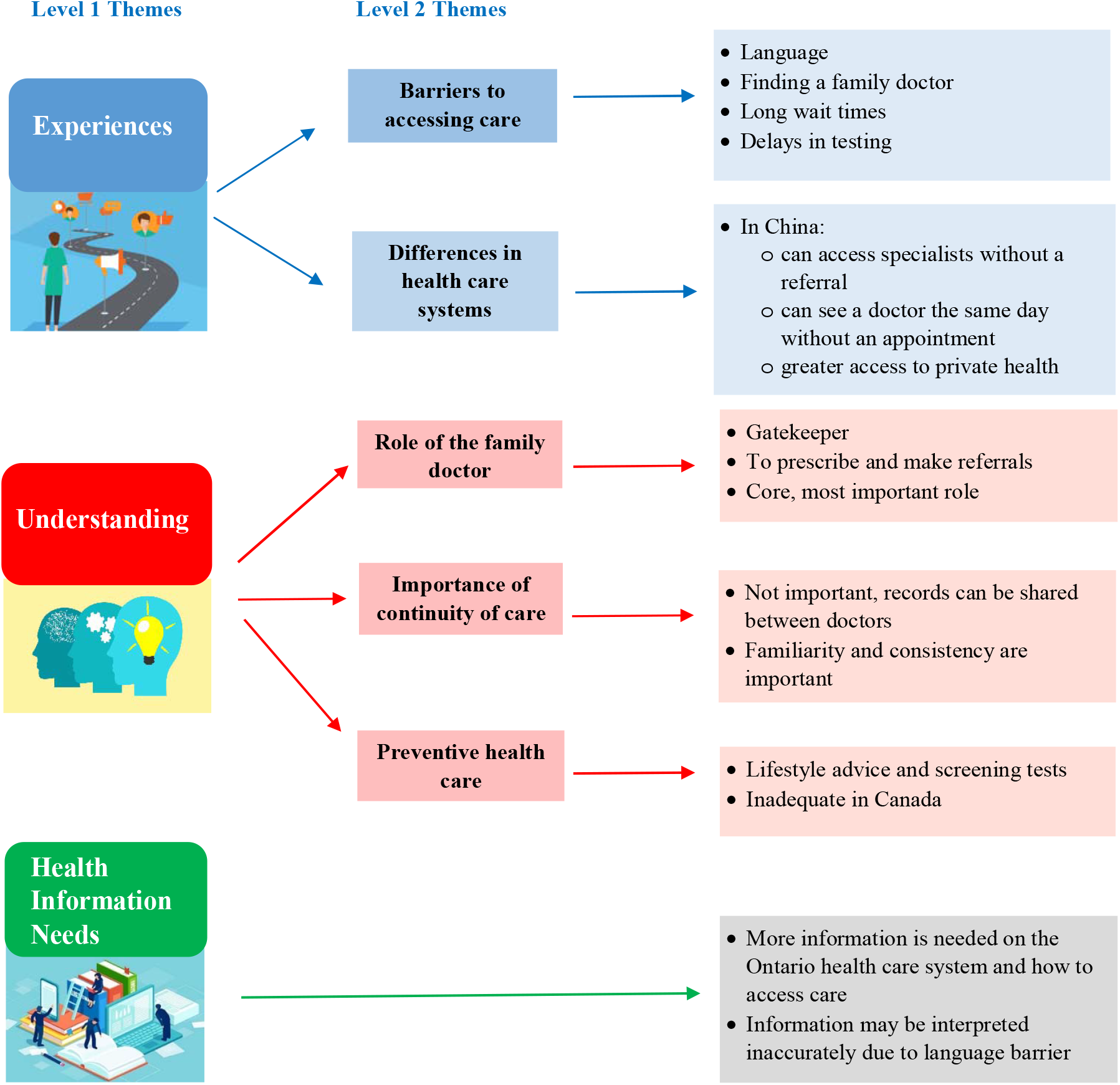
Thematic Map of Chinese Newcomers’ Perceptions of Primary Care.

### Barriers to Accessing Care

Participants described multiple challenges with accessing primary care in Ontario. Some identified difficulty finding a family doctor, especially one who spoke Mandarin. Others described lengthy wait times to see a family doctor or specialist, as well as perceived delays in obtaining investigations that were ordered by their physician.

> *“When you want to see a doctor, then it’s like the joke people make: ‘in Canada seeking medical care means waiting for things to self-heal*.*’ Maybe by the time you see them you are already recovered, or maybe you became worse and had to go to the emergency room*.*”* (Participant #9)
>
> *“We came here not long ago, and we are very uncomfortable with the completely different medical systems between the two countries. Sometimes when you want medical care, you must wait a long time to make an appointment with Ontario’s family doctors*.*”* (Participant #1)
>
> According to some participants, these barriers created a sense of anxiety and frustration. A few indicated that they go to the emergency department when they are unable to see their family doctor.

### Differences in Healthcare Systems

Participants commented on how differently the healthcare system is structured in Canada compared to China which could have significant implications for how care is accessed. Some participants expressed frustration that they could not obtain medication or tests that they felt were indicated or directly access specialists without a referral from a family physician. Others, however, felt that family physicians should determine the appropriateness of investigations and referrals.

> *“In Chinese hospitals, you can see any specialist and arrange testing of whatever organ you have problems with. But here, if the GP thinks you don’t need it, they won’t check it for you*.*”* (Participant #6)
>
> *“In China, you can check whatever you want, as long as you pay. But because healthcare is free here, there are more restrictions*.*”* (Participant #1)
>
> *“In China, you can go see a specialist in hospital immediately if you have a medical issue. If you have an urgent concern in Canada, you can only go to emergency*.*”* (Participant #4)

### Perceptions of the Role of the Family Doctor

Participants identified the family doctor as the gateway to medical care in Ontario. While some felt that the family doctor served an important role, others viewed this role narrowly – as limited to prescribing, ordering tests, or providing referrals. Some questioned the need for primary care suggesting that healthcare resources may be better spent by expanding access to specialists.

> *“For ordinary residents like me, the family doctor is the core role, the most important role. If you don’t need to go to hospital, your health problems are solved by your family doctor*.*”* (Participant #2)
>
> *“I don’t find him helpful. His role is only to prescribe medicine and make referrals*.*”* (Participant #1)
>
> *“I don’t understand why they fund GPs and not specialists. Specialists have better clinical acumen than GPs … I’d rather see a specialist because I trust their opinion more*.*”* (Participant #6)

### Importance of Continuity of Care

Participants had varying views regarding the importance of continuity of care. Some felt this could enhance care, others believed it was not important, and one participant thought that continuity is a hindrance to seeking specialist care.

> *“I think it’s very important [to see the same doctor over time], because the same doctor will know my situation the best…”* (Participant #4)
>
> *“I don’t think it’s important [to see the same doctor over time]. I know that the medical system has records which can be shared between doctors*.*”* (Participant #3)
>
> *“I think because the family doctor knows my husband for 20 years, and he knows that there’s no family history of any serious illness. So, he doesn’t feel necessary to refer him to see a specialist…”* (Participant #8)

### Perceptions of Preventive Healthcare

Participants generally thought that preventive healthcare was important, but they defined preventive healthcare as annual physical examinations and comprehensive investigations. Some were otherwise unsure of the meaning of the term ‘preventive healthcare’. Most participants believed that Canada offered inadequate preventive healthcare, especially compared with their previous experiences in China.

> *“Preventative care means that if I have any illness, the family doctor will give me some basic medical knowledge for my disease so that I know how to take care of my body. But it seems that I haven’t received this kind of preventative care here. Prevention is through physical checkups, but the physicals are not comprehensive*. (Participant #5)
>
> *“In China, women my age get gynecological examinations every year, and this is a comprehensive examination. But here, it is every 3 years. The things they check are also very rudimentary … If there is occult disease, it may not be detected through Canada’s screening*.*”* (Participant #1)

### Healthcare System Information Needs

Participants received information about the Ontario healthcare system from various sources including family, friends, internet, and community agencies. They generally wanted more information about how the Ontario healthcare system works and how to access care.

*“I don’t understand much about the services here* … *I’m not sure and I don’t know where to get this information*.*”* (Participant #7)

*“I would greatly appreciate if there was more information. Like how it works, what benefits people can get from family doctors*.*”* (Participant #10)

## Discussion

Newcomers in this study described experiences with healthcare in Ontario that are concordant with those described in previous Canadian research [1-10]. Like previous studies, they contrasted their experiences in Ontario with those in their country of origin. Most participants described healthcare in China as being superior to what they have experienced in Canada. A recent qualitative study of newcomers from various countries of origin explored experiences with healthcare in Regina, Saskatchewan [7]. Like our findings, participants felt that, compared to the care they received in Canada, healthcare in their country of origin was more supportive, usually delivered by a specialist, and involved more investigations [7].

This study also explored how newcomers from China perceived the role of the family doctor. Many of the participants viewed the family doctor as a gatekeeper, or even a barrier to perceived needed care. A similar finding was found in the Regina study [7]. Some of our participants stated that they lacked confidence in the family doctor’s ability and preferred instead to be able to access specialists directly. It is possible that this lack of trust in primary care services may relate to previous experiences in China. Chinese researchers have identified significant shortfalls in primary care in China stemming from insufficient training for primary care physicians [14]. This suggests that helping newcomers to understand the training involved in becoming a family doctor in Canada, and the integral role that they serve in the healthcare system, may improve the confidence and trust that newcomers have in family doctors.

Participants in our study felt that family doctors were reluctant to order tests and investigations primarily because of healthcare resource constraints. Although the Chinese government has recently implemented changes, the traditional fee for service primary care model may financially incentivize providers to order diagnostic tests and write prescriptions [14]. Having experienced healthcare interactions in China that are more likely to be accompanied by diagnostic tests and prescriptions, it is understandable that newcomers may perceive healthcare in Ontario to be inadequate if diagnostic tests are not performed and fewer prescriptions are written. Choosing Wisely Canada aims to decrease unnecessary testing and treatment not only to conserve healthcare resources, but also to reduce harm to patients [20]. Our findings highlight the importance of taking the time to explain to patients about evidence-based selection of diagnostic tests and the potential for inadvertent harm from unnecessary testing.

Participants had varying perspectives about the value of continuity of care. Studies have demonstrated that continuity of care is associated with a lower risk of hospitalization, reduced emergency department use, improved population health, enhanced patient satisfaction, and reduced healthcare costs [21-22]. Patients with multimorbidity are more likely to value continuity of care, even for acute visits, compared to healthier patients [23]. Our participants were young, with few chronic diseases, which may have influenced their perspectives on the value of continuity of care. It cannot be expected that newcomers to Canada will intuitively appreciate the benefits of continuity of care.

Understandably, patients may prioritize ease of access over continuity, especially when systemic factors often limit access.

There is an opportunity to help orient newcomers to the Ontario healthcare system by explaining the benefits of continuity of care and preventive healthcare, and guiding appropriate use of available healthcare options (e.g. family physician, walk-in-clinic, emergency room). Family physicians can play an important role in this process however, newcomers usually do not have a family physician upon arrival and may be unaware of the importance of finding a primary care provider. Participants in this study reported learning about the Canadian healthcare system from various sources including family, friends, government websites, and community agencies, highlighting the absence of a systematic approach. A recent scoping review of immigrants’ experience of healthcare access in Canada recommended developing a culturally sensitive community navigator program to support healthcare system navigation [2]. Strengthening intersectoral collaboration between primary care teams and other community organizations (such as settlement centres) could further enhance health system orientation for newcomers.

## Limitations

Participants were recruited through a settlement agency and may therefore have been better informed about the healthcare system compared to newcomers who did not access such services. Most participants did not have chronic health conditions which may have influenced their perspectives on continuity of care. Participants were well educated and likely had access to private healthcare in China, although this was not explicitly assessed. Greater diversity in socioeconomic background or health status may have yielded different findings. While the general good health of participants may represent a study limitation, it aligns with the well-documented “healthy immigrant effect” whereby, upon arrival, newcomers are generally healthier than the native-born population [23-24]. Finally, China, like Canada, is a vast and diverse country and thus participants’ health experiences may have varied considerably before coming to Canada.

### New Contribution to the Literature

This study adds to the literature by exploring newcomers’ understanding of preventive healthcare and continuity of care, two core tenets of primary care in Canada. Participants believed that Canadian preventive healthcare was rudimentary and did not entail sufficient testing. Newcomers may appreciate the opportunity to learn more about the evidence-based processes that guide preventive healthcare programs in Canada. Although this study focused on the perceptions of newcomers from mainland China, there are broader implications for newcomers from other countries. Similarly, expectations and perceptions of primary care in their new home are likely influenced by experiences with healthcare in their country of origin. It is important to understand and acknowledge these differences as they may influence how people seek care in their new home.

There is a clear need to improve newcomer orientation to the role of primary care in Ontario. Newcomers from China identified barriers to accessing care in the Canadian healthcare system. Enhancing newcomers’ understanding of the Canadian healthcare system may help reduce these barriers and improve system navigation. Future research should explore experiences and perceptions of newcomers from other countries. Meanwhile, there is significant potential for collaboration between healthcare providers and community agencies to support newcomer orientation to primary care.

## Data Availability

All data produced in the present work are contained in the manuscript.

## Acknowledgements

We thank the Centre for Immigrant and Community Services for their support and assistance with recruitment for
this study.

